# Interactive effects of genotype with prenatal stress on DNA methylation at birth

**DOI:** 10.1101/2024.11.20.24317575

**Authors:** Rosa H. Mulder, Vilte Baltramonaityte, Serena Defina, Katerina Trajanoska, Matthew Suderman, Emanuel Schwarz, Marco P. M. Boks, Esther Walton, Charlotte A. M. Cecil, Janine F. Felix

## Abstract

Intrauterine stress exposure is associated with offspring health. DNA methylation (DNAm) is as a putative underlying mechanism, but large population-based studies reported limited associations between prenatal stress and DNAm. Recent research has shown that environmental factors *in interaction* with genetic variants are better predictors of DNAm than environment or genotype alone. We investigated whether interactions of maternal prenatal stress with genetic variants are associated with DNAm at birth. We examined 2,963 mother-child pairs from the population-based Generation R Study and Avon Longitudinal Study of Parents and Children, using a harmonized, comprehensive cumulative prenatal stress measure. We tested genome-wide genotype-by-prenatal stress interactions on epigenome-wide DNAm (GxEmodel), and models including only genetic variants (Gmodel) or prenatal stress (Emodel) as predictors. Follow-up analyses included Gene Ontology analyses and mediation analyses of prenatal alcohol intake, smoking, gestational age, and birth weight. We report two independent gene-by-prenatal-stress interactions on DNAm after multiple testing correction, including five genetic variants in *CHD2* and *ORC5,* and two DNAm sites in *EPPK1*. By comparison, the Gmodel showed 691,202 associations and the Emodel showed three associations in genes *AHRR, GFI1*, and *MYO1G*, which could largely explained by prenatal smoking. Genes linked to suggestive GxEmodel results were often involved in neuronal development. Our results provide some support of interaction of prenatal stress with the child’s genome on DNAm of genes related to neuronal development. These results do not confirm the notion that gene-by-environment interaction models show more associations with DNAm compared to genes or the environment studied in isolation.

## Introduction

*In utero* stress exposure has been associated with adverse offspring mental and physical health outcomes, including internalizing symptoms^1^, adiposity^2^, asthma, and allergies^3^, and has been hypothesized to put children in a disadvantaged position from early life onwards. Differential DNA methylation (DNAm) has been suggested as a putative mechanism underlying these associations, as DNAm has been linked to prenatal exposures such as maternal smoking^4^, postnatal psycho-social stress^5^, and to child outcomes such as body mass index^6^, asthma^7^ and cortisol reactivity^8^. Several multi-cohort studies have probed epigenome-wide associations of maternal prenatal stress with offspring DNAm, with varying results^9–11^. The largest study to date, including 5,496 children from 12 cohorts, reported limited associations for DNAm sites located in genes that have been implicated in neurodegeneration, immune and cellular functions, and epigenetic regulation^10^.

A growing body of research, however, shows that environmental factors *in interaction with* genetic variants are better predictors of DNAm than environmental factors or genetic variants alone. For example, Teh, Pan ^12^ studied genome-wide interactions of 19 prenatal factors, including gestational age, maternal smoking and maternal depression, on highly variable neonatal DNAm sites. For 75% of the sites, DNAm was better predicted by the interaction between genotype and the environment than by either genotype or environment alone. Environment-only was never the best predictor of DNAm in that study. Similarly, a study by Czamara, Eraslan ^13^ in four cohorts, examining 10 prenatal factors, showed that gene-environment interactions best predicted DNAm in variably methylated regions in 38-60% of analyses, while genotype-only models were best in 11-30% and environment-only models were best in only <1-4%. However, while these studies analyzed which *type* of model worked best, they did not aim to identify specific genetic variants, environmental variables or DNAm sites. Knowing which genetic variants interact with prenatal stress in relation to DNA methylation would help to better understand the biological pathways underlying the gene-environment effects on health.

We therefore aimed to study genome-wide interactions between genetic variants and cumulative prenatal stress in relation to epigenome-wide DNAm at birth. We also aimed to test the hypothesis that DNAm is better predicted by the interaction of genetic variants and stress than by either factor alone. We used a comprehensive cumulative measure of psycho-social maternal stress during pregnancy, which has previously been related to suboptimal neurodevelopmental, mental, and cardiovascular outcomes^14–16^. We meta-analyzed data from two population-based cohorts, the Generation R Study in the Netherlands (Generation R) and the Avon Longitudinal Study of Children and Parents (ALSPAC) in the United Kingdom and followed up associations to study unique stress domain contributions, as well as running mediation analyses of maternal prenatal smoking, alcohol use, gestational age and birth weight. Lastly, we performed enrichment analyses to gain insight into potential biological pathways.

## Methods

### Setting

We used three non-overlapping datasets from two prospective population-based cohorts: two datasets from Generation R and a third dataset from ALSPAC.

In the Generation R Study, pregnant women residing in the study area of Rotterdam in the Netherlands with an expected delivery date between April 2002 and January 2006 were invited to participate in the study^17^. The Generation R Study is conducted in accordance with the World Medical Association Declaration of Helsinki and has been approved by the Medical Ethics Committee of Erasmus MC, Rotterdam. Informed consent was obtained for all participants.

In ALSPAC, pregnant women resident in Avon, UK with expected dates of delivery between 1^st^ April 1991 and 31^st^ December 1992 were invited to take part in the study^18, 19^. The ALSPAC website contains details of all the data that are available through a fully searchable data dictionary and variable search tool (http://www.bristol.ac.uk/alspac/researchers/our-data/). Ethical approval for the ALSPAC study was obtained from the ALSPAC Ethics and Law Committee and the Local Research Ethics Committees. Informed consent for the use of data collected via questionnaires and clinics was obtained from participants following the recommendations of the ALSPAC Ethics and Law Committee at the time. Consent for biological samples has been collected in accordance with the Human Tissue Act (2004).

### Study Population

The full selection procedure is described in the **Supplemental Information**. In Generation R, 9,778 pregnant mothers gave birth to 9,749 live-born children and in ALSPAC, the initial number of pregnancies enrolled was 14,541. Participants were selected based on availability of genetic data (*n*_Generation R_=7,502; *n*_ALSPAC_=8,797), as well as cumulative prenatal stress information (*n*_Generation R_=5,684; *n*_ALSPAC_= 7,483), and DNAm data as measured with the Infinium HumanMethylation450 BeadChip (Illumina Inc., San Diego, CA) in Generation R (*GENR 450K*) and ALSPAC (*ALSPAC 450K*) or with the Infinium MethylationEPIC v1.0 Beadchip in Generation R (*GENR EPIC*) (*n*_GENR 450K_=1,231; *n*_GENR EPIC_=986; *n*_ALSPAC_=793). Additionally, one of each pair of children with cryptic relatedness (IBD>0.15) were removed, based on data availability or otherwise randomly. As a result, *GENR 450K* included 1,224 children, *GENR EPIC* included 949, and *ALSPAC 450K* included 790 – for a total of 2,963 children.

### Genotyping

In the Generation R Study, children were genotyped with the Illumina HumanHap 610 or 660 quad chips. A full description has been published previously^20^. Data were imputed to the 1000 genomes reference panel (Phase 1 version 3). Phasing was done using MACH software, and imputation using Minimac software. The ALSPAC children have been genotyped with the Illumina HumanHap 550 quad chip^21^. The data were imputed to a phased version of the 1000 genomes references panel (Phase 1 version 3) from the Impute2 reference data repository.

In all (sub-)cohorts, we used best-guess genotypes. Quality control was done with PLINK 1.90^22^. Autosomal variants were selected and variants with SNP call rates of <95%, with evidence for violation of Hardy-Weinberg equilibrium (*p*<1×10^-07^), with a minor allele frequency <5%, or with low imputation quality (Rsq<0.3 in Generation R and info scores <0.8 in ALSPAC, according to local practices^20, 21^) were removed. Insertions, deletions, and multi-allelic positions were also removed. Samples were excluded in the case of sex mismatches, minimal or excessive heterozygosity, or a sample call rate of <97.5%.

This quality control procedure resulted in 5,584,862 SNPs in *GENR 450K*; 5,627,497 SNPs in *GENR EPIC;* and 5,797,754 SNPs in *ALSPAC 450K.* To reduce the multiple testing burden, SNPs were pruned based on linkage disequilibrium and haplotype blocks (window size=50 SNPs, step size=5 SNPs, VIF=2) in the largest sub-cohort, *GENR 450K,* which resulted in 447,713 SNPs. Of these, a final set of 374,152 SNPs was common to all three (sub-)cohorts.

### Cumulative prenatal stress

The cumulative prenatal stress score was computed from ∼50 stress-related items measured during pregnancy. The full item list and a detailed description of the score calculation can be found elsewhere (https://github.com/SereDef/cumulative-ELS-score^15, 16^). Briefly, in order to maximize data harmonization across cohorts, stress items were selected (based on closest item-similarity), dichotomized (0=no risk; 1=risk) and assigned to one of four stress domains: life events (e.g. death of a relative), contextual risk (e.g. financial problems), personal stress (e.g. depression), and interpersonal stress (e.g. family conflict). Stress domain scores (ranging from 0 to 1) were then computed by averaging items within each domain. A total prenatal stress score was obtained by summing all domain scores (range: 0 to 4). Individuals with >50% of all stress items missing were excluded. Missing data were imputed at the individual item level using predictive mean matching with 60 iterations, as implemented by the *mice* package^23^ in R version 4.0^24^. Within the selected samples of each (sub-)cohort, cumulative prenatal stress scores were standardized. To reduce the influence of extreme outliers, we winsorized values outside the range of (25^th^ percentile - 3*interquartile range (IQR)) to (75^th^ percentile + 3*IQR).

### DNA methylation

For both cohorts, DNA extracted from cord blood was bisulfite converted. Samples were processed with the Illumina Infinium HumanMethylation450 BeadChip (Illumina Inc., San Diego, CA) in *GENR 450K* and *ALSPAC 450K* and with the Infinium MethylationEPIC v1.0 Beadchip in *GENR EPIC*.

In *GENR 450K* and *GENR EPIC*, the CPACOR workflow^25^ was applied for quality control. Arrays with observed technical problems such as failed bisulfite conversion, hybridization or extension as well as arrays with a sex mismatch were removed. Arrays with a call rate >95% per sample were carried forward into normalization.

In *ALSPAC 450K*, quality control was done using the *meffil* package^26^ in R version 3.4.3. Samples with mismatched genotypes, mismatched sex, incorrect relatedness, low concordance with samples collected at other time points, extreme dye bias and poor probe detection were removed and carried before normalization.

In order to minimize cohort effects, the data from *GENR 450K and ALSPAC 450K* have been previously normalized as a single dataset ^27^ and data from *GENR EPIC* were normalized using the same procedure. Functional normalization was performed (using 10 control probe principle components with slide included as a random effect) with the *meffil* package in R^26^. Probes were excluded if they had a detection *p*>0.01 or low bead count (<3) in >10% of the samples. In total, 472,450 autosomal methylation sites (CpGs) passed these quality control filters in *GENR 450K* and *ALSPAC 450K*. To reduce the computational burden of the genome-wide analyses on the methylome, only probes that were previously identified as having epigenome-wide significant (*p*<1×10^-7^) inter-individual variation in DNAm at birth in these cohorts were carried forward into analyses, leaving 100,687 CpGs^27^. Of these, cross-reactive probes (*n*=14,451 CpGs) were removed^28^. Last, only probes present and passing quality control on both array types were selected, resulting in a final set of 86,236 CpGs. DNAm levels were represented as beta values, indicating the ratio of methylated signal relative to the sum of methylated and unmethylated signal per CpG. To reduce the influence of extreme outlying values, beta values of each CpG outside the range of (25^th^ percentile - 3*interquartile range (IQR)) to (75^th^ percentile + 3*IQR) were winsorized.

### Covariates

All models were adjusted for sex of the child as determined at birth, the first 5 genetic principle components, estimated blood cell composition (CD4+ T-lymphocytes, CD8+ T-lymphocytes, natural killer cells, B-lymphocytes, monocytes, granulocytes, and nucleated red blood cells) as based on a cord blood reference panel^29^, and DNAm batch effects (25 sample plates in *GENR 450K,* 12 sample plates in *GENR EPIC*, and 20 surrogate variables in *ALSPAC 450K*^26, 30^).

### Statistical analyses

Analyses were performed with an adapted version^16^ of the *GEM* software package^31^ in R^24^ version 4.0.5. This package applies large matrix operations allowing for fast analysis of genome-wide SNPs and CpGs and enabling us to test the following models: (i) a *GxEmodel* – our primary model of interest – in which the interaction effect of each SNP with cumulative prenatal stress was iteratively regressed on DNAm at each CpG site. For comparative purposes, we also tested (ii) a *Gmodel*, in which each SNP was iteratively regressed on DNAm at each CpG site, and (iii) an *Emodel*, in which cumulative prenatal stress was iteratively regressed on DNAm at each CpG site. A dominant model was applied to the GxEmodel and Gmodel, meaning that heterozygous and homozygous minor genotypes were contrasted against homozygous major genotypes, in order to make the models more robust against outlying values. The three models were performed in each (sub-)cohort separately, and results were meta-analyzed using inverse-variance weighted fixed effects with METAL^32^. The significance threshold of the meta-analyses was Bonferroni-corrected for the number of tests. For the GxEmodel and Gmodel (*n*_SNPs_=374,152, *n*_CpGs_=86,236: 32,265,371,872 tests) the threshold was set to *p*<1.55×10^-12^, and for the Emodel (86,236 tests), the threshold was set to *p*<5.80×10^-07^. To assess heterogeneity between (sub-)cohort results, the I^2^ statistic was used, with 75% taken as indication of considerable heterogeneity^33^.

### Model comparisons

Earlier studies tested SNP-by-environment interactions with CpGs only among SNPs or CpGs for which an association was found in a genotype-only. We tested whether SNPs/CpGs that showed suggestive associations in the Gmodel or Emodel had a higher chance of being part of a suggestive association in the GxEmodel. Associations in the Gmodel and the GxEmodel with *p*<5×10^-08^ (i.e. genome-wide threshold), and associations in the Emodel with *p*<1×10^-05^ were considered suggestive. We performed enrichment analyses comparing suggestive versus non-suggestive SNPs and CpGs using Fisher’s exact tests (significance threshold: *p*<0.05).

As a sensitivity analysis, we performed similar enrichment analyses to assess if suggestive findings in the GxEmodel were more or less likely to have been identified as (SNPs) or associated with (CpGs) a methylation quantitative trait locus (meQTL) in an earlier large-scale study of DNAm at different ages^34^.

### Follow-up analyses

Multiple follow-up analyses were performed. Since the Gmodel has been tested extensively previously in search of methylation quantitative trait loci (meQTLs)^21, 34^, we only followed-up results from the GxEmodel and Emodel. First, we looked up significant CpGs in the GxEmodel and/or Emodel in the EWAS Catalog for previously reported associations^35^. Second, we looked up genes annotated to significant SNPs and CpGs in the GxEmodel and/or Emodel via phenome-wide association studies (PheWASs) using the online GWAS Atlas tool (https://atlas.ctglab.nl/PheWAS)^36^, including 4,756 GWASs, using a Bonferroni corrected *p*<1.05×10^-05^. Annotation of SNPs and CpGs was performed using ANNOVAR linking variants reported in the 1000 Genomes Project and SNPdb^37^ and the Illumina HumanMethylation450 v1.2 Manifest (Illumina Inc.), respectively. Third, significant associations in the GxEmodel (*p*<1.55×10^-12^) and/or Emodel (*p*<5.80×10^-07^) were followed up with linear regressions containing the effects of the four stress domains (life events, contextual risk, personal stress, and interpersonal stress) in one model to identify unique associations of each of these stress types, independent of the other types. Here, associations with *p*<0.05 were interpreted as a unique contribution to the GxE association on DNAm for that stressor. Fourth, significant associations in the GxEmodel and/or Emodel were tested for potential mediation of prenatal stress effects on DNAm by maternal prenatal smoking, maternal prenatal alcohol intake, gestational age, and birth weight (each modeled separately), using the Lavaan package in R^38^. Mediation was deemed to be significant if the AB path (predictor -> mediator, mediator -> outcome) has a *p*-value below a Bonferroni-corrected threshold of 0.0125 (corrected for the number of mediators). Last, functional enrichment analysis of associated biological pathways was performed with Gene Ontology for models for which results could not be explained by a mediator. Genes annotated to suggestive unique SNPs and CpGs were interrogated using the GOfuncR package^39^ in R, using the built in family-wise error rate correction for multiple testing.

### Mediators

In Generation R, mothers reported on prenatal tobacco smoking and alcohol consumption via questionnaires in the first, second, and third trimester. In ALSPAC, mothers reported via questionnaires on tobacco smoking in the second and third trimester and on alcohol consumption in the first and second trimester. For both cohorts, gestational age at birth was determined using fetal ultrasound examinations or last menstrual period, and birth weight was obtained from midwife and hospital registries.

## Results

*GENR 450K*, *GENR EPIC*, and *ALSPAC 450K* included 49.9%, 47.7%, and 49.1% boys, and mothers were 32.2, 32.0, and 29.7 years old at birth, respectively. After winsorizing, mean cumulative prenatal stress scores were 0.36 (SD=0.28, min=0.00, max=1.48), 0.44 (SD=0.35, min=0.00, max=1.48), and 0.51 (SD=0.28, min=0.00, max=1.69), respectively, with a theoretical maximum score of 4 (**Supplemental Figure 1**).

### GxEmodel: SNP by prenatal stress interactions and DNA methylation

Five SNP-by-prenatal-stress interactions on DNAm were identified after Bonferroni correction, including five unique SNPs and two unique CpGs. Firstly, an association in cis of rs12901653 in CHD2 in interaction with cumulative prenatal stress was found for DNAm at a nearby (64862 bp) cg24317086 (B=-0.026, SE=0.003, p=4.07×10^-16^), for which CHASERR, or CHD2 Adjacent Suppressive Regulatory RNA is the nearest gene. Secondly, trans-associations of 4 SNPs in or near ORC5 in interaction with cumulative prenatal stress were found for DNAm at cg06592260, which is located in EPPK1 (rs7642426: B=-0.013, SE=0.002, p=1.12×10^-13^; rs10279675: B=-0.012, SE=0.002, p=2.76×10^-13^; rs2188287: B=-0.013, SE=0.002, p=1.85×10^-13^; rs10251976: B=-0.012, SE=0.002, p=7.32×10^-13^). As these SNPs correlated highly (>0.9 in all [sub-]cohorts), these were not independent. Results are depicted in Table 2 and Figure 1. For all associations except the interaction of rs12901653 with cumulative prenatal stress on cg24317086, heterogeneity between (sub-)cohorts was low (I^2^ =0.0). Rs12901653 showed considerable heterogeneity (I =93.5), as associations for GENR 450K and GENR EPIC were negative, whereas it was positive (although not significant) for ALSPAC 450K (forest plot in Supplemental Figure 2).

**Figure 1.**
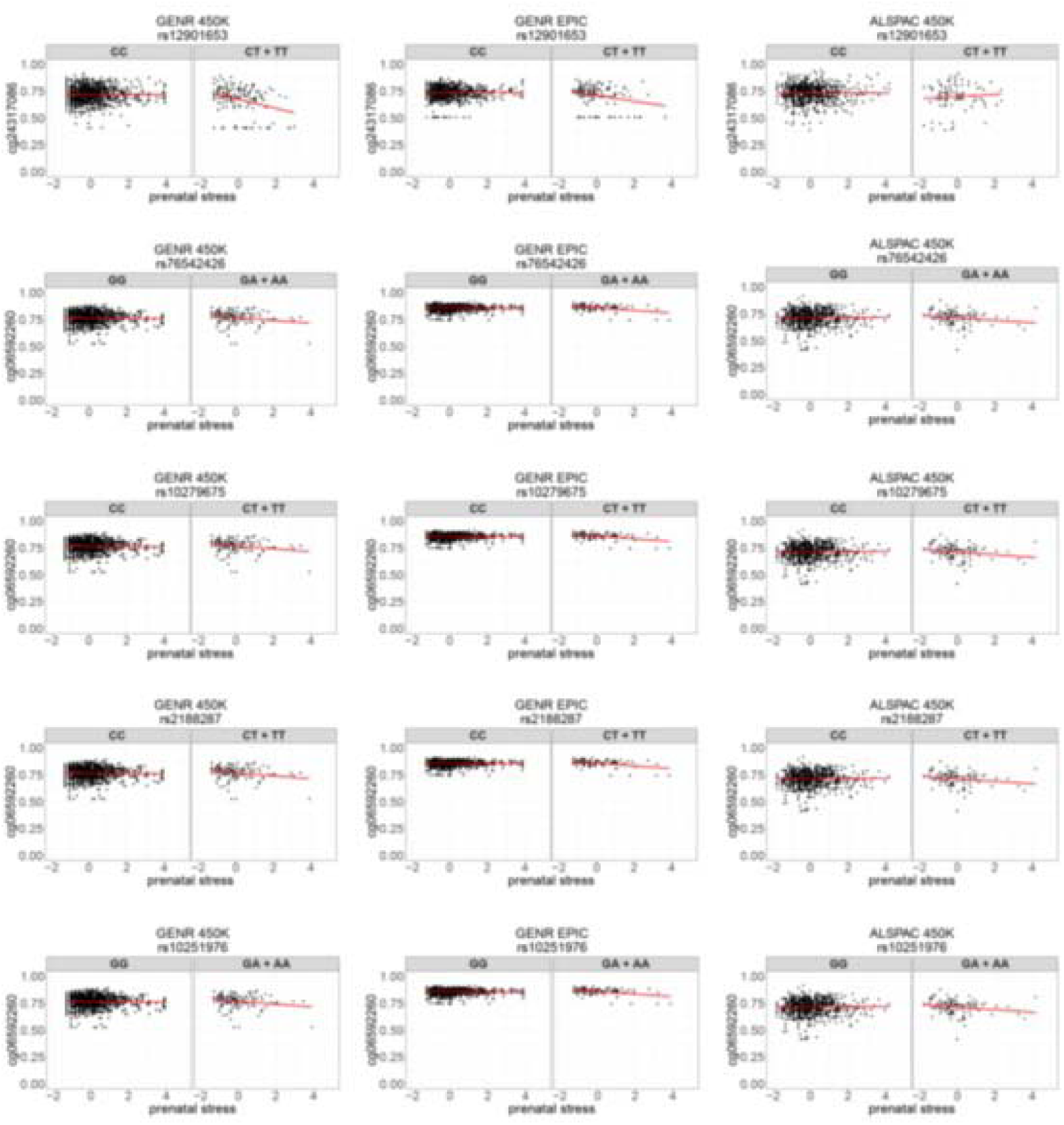
Scatterplots of genome- and epigenome-wide associations of SNP-by-prenatal-stress interactions and DNA methylation.

**Table 1.**
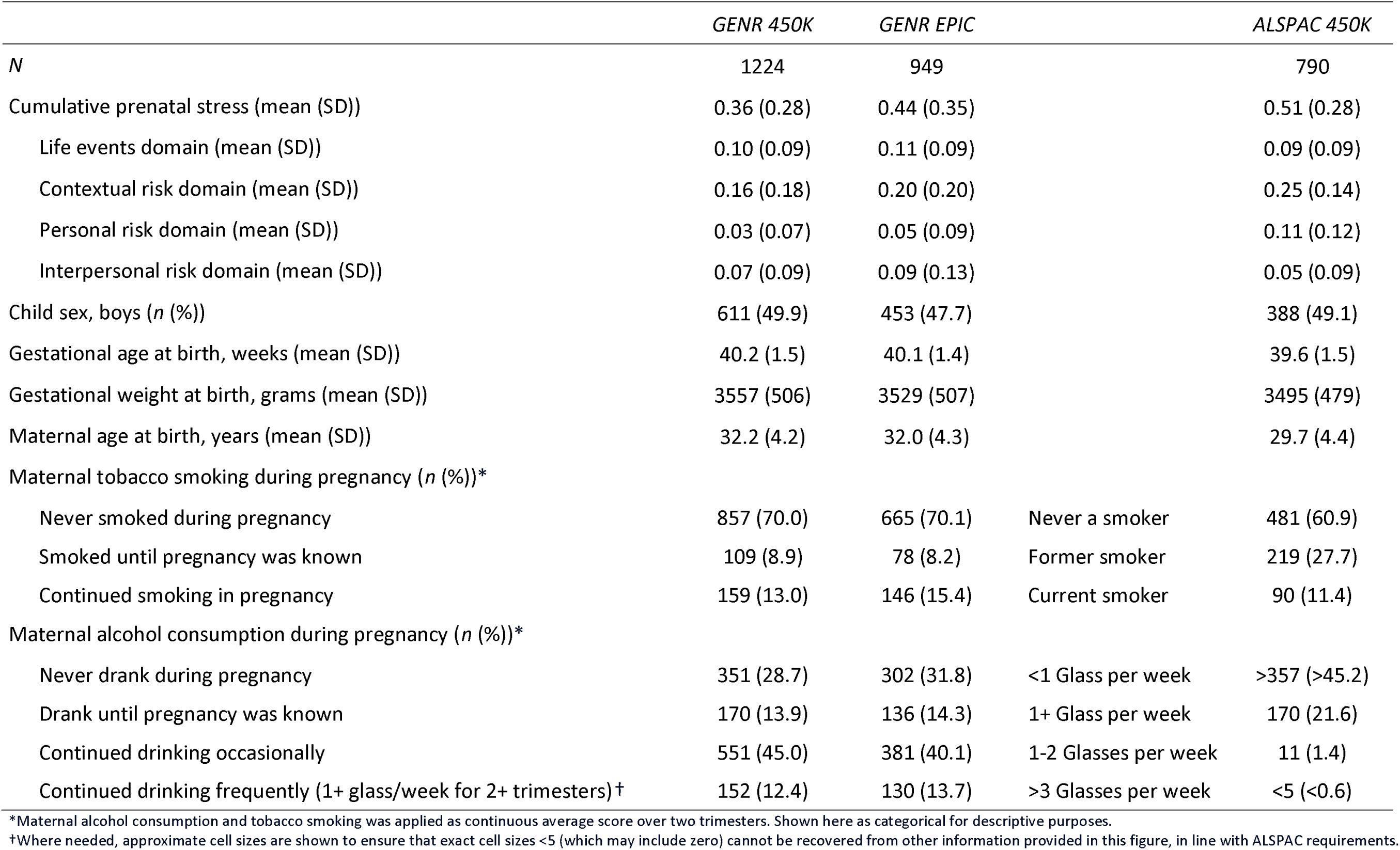
Sample characteristics.

**Table 2.**
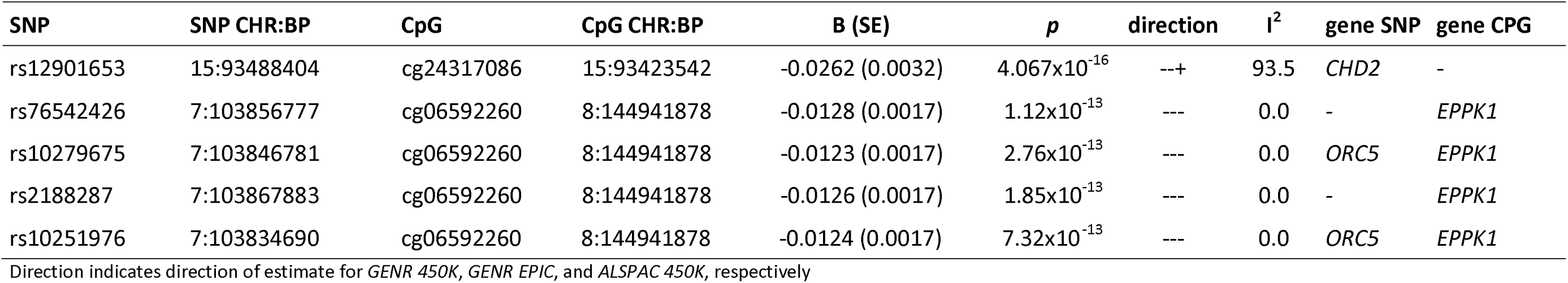
Genome- and epigenome-wide associations of SNP by cumulative prenatal stress interactions and DNA methylation.

| SNP | SNP CHR:BP | CpG | CpG CHR:BP | B (SE) | *p* | direction | I<sup>2</sup> | gene SNP | gene CPG |
| --- | --- | --- | --- | --- | --- | --- | --- | --- | --- |
| rs12901653 | 15:93488404 | cg24317086 | 15:93423542 | -0.0262 (0.0032) | 4.067x10<sup>-16</sup> | --+ | 93.5 | *CHD2* | - |
| rs76542426 | 7:103856777 | cg06592260 | 8:144941878 | -0.0128 (0.0017) | 1.12x10<sup>-13</sup> | --- | 0.0 | - | *EPPK1* |
| rs10279675 | 7:103846781 | cg06592260 | 8:144941878 | -0.0123 (0.0017) | 2.76x10<sup>-13</sup> | --- | 0.0 | *ORC5* | *EPPK1* |
| rs2188287 | 7:103867883 | cg06592260 | 8:144941878 | -0.0126 (0.0017) | 1.85x10<sup>-13</sup> | --- | 0.0 | - | *EPPK1* |
| rs10251976 | 7:103834690 | cg06592260 | 8:144941878 | -0.0124 (0.0017) | 7.32x10<sup>-13</sup> | --- | 0.0 | *ORC5* | *EPPK1* |

Direction indicates direction of estimate for *GENR 450K*, *GENR EPIC*, and *ALSPAC 450K*, respectively

### Gmodel: SNPs and DNA methylation

In the Gmodel, after Bonferroni correction, we found 691,202 associations between SNPs and DNAm, including 181,133 unique SNPs and 54,809 unique CpGs. As such, nearly half of all investigated SNPs (48%) could be marked as meQTLs, and more than half (59%) of the examined CpGs are under genetic control. In these results we find evidence of both polygenicity, i.e. multiple SNPs affecting the same CpG, as well as pleiotropy, i.e. the same SNP affecting multiple CpGs. Furthermore, 91% of SNP-CpG associations were in cis, 9% were in trans (distance of >1,000,000 bp). For 28% of associations, heterogeneity between (sub-)cohort results was considerable (I^2^ >75).

### Emodel: Cumulative prenatal stress and DNA methylation

Three DNAm sites at birth were associated with exposure to cumulative prenatal stress after Bonferroni correction, including cg05575921 (B=-0.009, SE=0.001, p=3.81×10^-18^), located in AHRR, cg09935388 (B=-0.016, SE=0.002, p=2.79×10^-11^) in GFI1, and cg04180046 (B=-0.007, SE=0.001, p=6.73×10^-08^) in MYO1G (Table 3; Figure 2). For cg05575921, there was considerable heterogeneity between (sub-)cohort results (I^2^=78.1), for cg09935388 and cg04180046 no heterogeneity was detected (I^2^=0.0; forest plot in Supplemental Figure 3).

**Figure 2.**
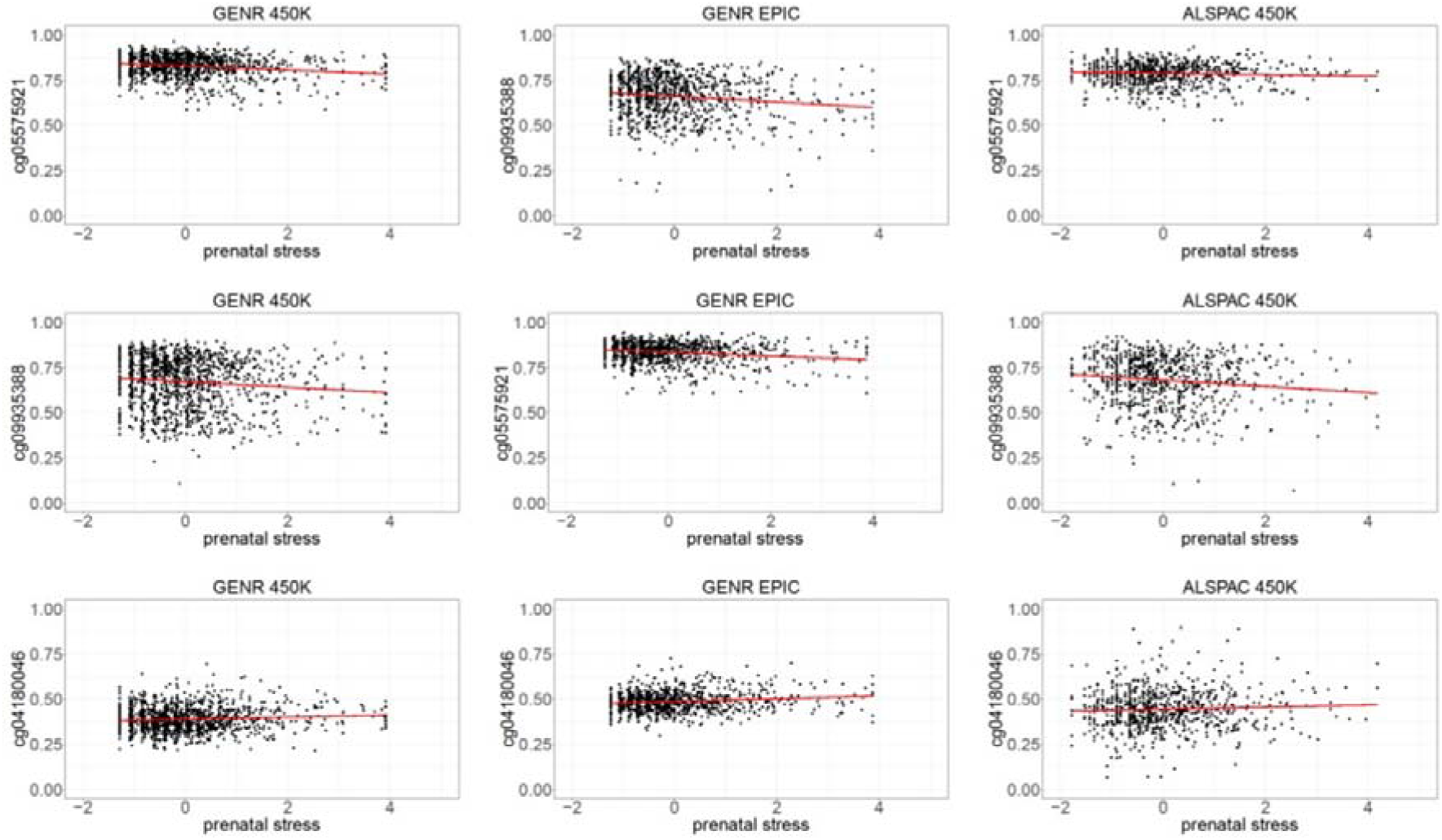
Scatterplots of epigenome-wide associations of cumulative prenatal stress and DNA methylation.

**Table 3.** Epigenome-wide associations of cumulative prenatal stress and DNA methylation.

| CpG | CHR:BP | B (SE) | *p* | direction | I<sup>2</sup> | gene |
| --- | --- | --- | --- | --- | --- | --- |
| cg05575921 | 5:373378 | -0.0086 (0.0010) | 3.810x10<sup>-18</sup> | --- | 78.1 | *AHRR* |
| cg09935388 | 1:92947588 | -0.0161 (0.0024) | 2.788x10<sup>-11</sup> | --- | 0.0 | *GFI1* |
| cg04180046 | 7:45002736 | 0.0070 (0.0013) | 6.732x10<sup>-08</sup> | +++ | 0.0 | *MYO1G* |

Direction indicates direction of estimate for *GENR 450K*, *GENR EPIC*, and *ALSPAC 450K*, respectively

### Enrichments of main effect model associations in GxEmodel

Suggestive SNPs and CpGs in the GxEmodel (3,248 SNPs and 2,613 CpGs) and Gmodel (223,254 SNPs and 62,826 CpGs) were compared to test whether a suggestive association in the Gmodel increased the chance of a suggestive association in the GxEmodel. This did not seem to be the case, as suggestive SNPs in the Gmodel were as likely to have been identified in the GxEmodel as other SNPs were (1% vs 1%; OR=0.9 [95% CI=0.9-1.0], *p*=0.13). Similarly, suggestive CpGs in the Gmodel were as likely to be identified in the GxEmodel as other CpGs were (3% vs 3%; OR=1.0 [95% CI=0.9-1.1], *p*=0.74). As a sensitivity analysis, we also checked for enrichment of SNPs and CpGs associated with meQTLs identified by others^34^, and similarly found that suggestive SNPs and CpGs in the GxEmodel were as, or even less, likely to have been linked to an meQTL, whereas suggestive hits in the Gmodel were more likely to have previously been linked to an meQTL (**Supplemental Results S1**).

Furthermore, suggestive CpGs in the Emodel were as likely to have been identified as a suggestive CpG in the GxEmodel (7%) as other CpGs were (3%; OR=2.5 [95% CI=0.1-16.4], *p*=0.35), although it should be noted that the number of suggestive findings for the Emodel was low.

### CpG look-ups

From the GxEmodel, variation at cg24317086 has been previously associated with gestational age^40^, age in childhood^27, 41^, tissue type^42^, Down syndrome^43^, and C-reactive protein levels^44^. Variation at cg06592260 has been associated with age in childhood^27^ and tissue type^42^.

Full results for the lookup of the previously reported EWAS associations for the three CpGs found in the Emodel can be found in **Supplemental Table 1.** In brief, variation at cg05575921, cg09935388, and cg04180046 was related to maternal smoking during pregnancy with reported associations stemming from 6, 8 and 9 studies, respectively, and to smoking behavior (not in pregnancy) with reported associations in 27, 19, and 10 studies, respectively. Other associations were found, among others, for age in childhood^27^, tissue type^42^, alcohol consumption^45–47^, maternal educational attainment during pregnancy^48^, lung function^49–52^ and post-traumatic stress syndrome^53,54^.

### Annotated gene look-up

The full results of the PheWASs are depicted in **Supplemental Figures 4 to 9.** In brief, genetic variants at *CHD2* were related to use of sun/UV protection, resting heart rate, free thyroxine levels, educational attainment, measures of body composition, uric acid levels, processed meat intake, pork intake, napping during the day, (standing) height, and schizophrenia. Genetic variants at *ORC5* have been related to risky behaviors, left and right entorhinal cortex volume, and drinking behavior. Genetic variants in or close to *EPPK1* (annotated to several CpGs of the significant GxEmodels) have been related to resting heart rate, skin tanning, body composition measures, and height.

Genetic variants at *AHRR* were related to skin colour, hair colour, male balding patterns, ulcerative colitis, hematocrit, hemoglobin, aspartate, fat measures, and height. Genetic variants annotated to *GFI1* were related to coronary artery disease, white blood cell measures, fat measures, multiple sclerosis, being a morning person, height, lung function, and asthma, eczema, and allergy related measures. Genetic variants at *MYO1G* were related to thyroid function, white blood cell measures, and height.

### Stress-domain-specific results

In the GxEmodel, none of the individual stress domains (life events, contextual risk, personal risk, interpersonal risk) provided a unique SNP-by-prenatal-stress contribution (*p*<0.05) to the association with DNAm, over and above co-occurring domains (**Supplemental Table 2**). In the Emodel, contextual risk provided a unique contribution to DNAm at cg05575921 (*AHRR*, B=-0.008, SE=0.001, *p*=4.42x^-14^), cg09935388 (*GFI1*, B=-0.015, SE=0.003, *p*=6.18×10^-09^), and cg04180046 (*MYO1G*, B=0.005, SE=0.001, *p*=7.36×10^-05^). In addition, interpersonal risk provided a unique contribution to cg05575921 (B=-0.003, SE=0.001, *p*=7.53×10^-03^) and cg09935388 (B=-0.007, SE=0.003, *p*=5.44×10^-03^). Life events and personal risk did not provide unique contributions in the significant Emodel associations (**Supplemental Table 3**).

### Mediation

In the GxEmodel, none of the significant SNP-by-prenatal-stress associations with DNAm were mediated by maternal tobacco smoking or alcohol consumption during pregnancy, gestational age, or birth weight. In the Emodel, all three cumulative prenatal stress associations with DNAm were mediated by maternal prenatal smoking (cg05575921: B_indirect_=-0.007, 95% CI=-0.008;-0.006, *p*=4.17×10^-60^; cg09935388: B_indirect_=-0.010, 95% CI=-0.011;-0.008, *p*=3.21×10^-30^; cg04180046: B_indirect_=-0.006, 95% CI=-0.005;-0.007, *p*=8.94×10^-40^), and not by any of the other mediators (**Figure 3**).

**Figure 3.**
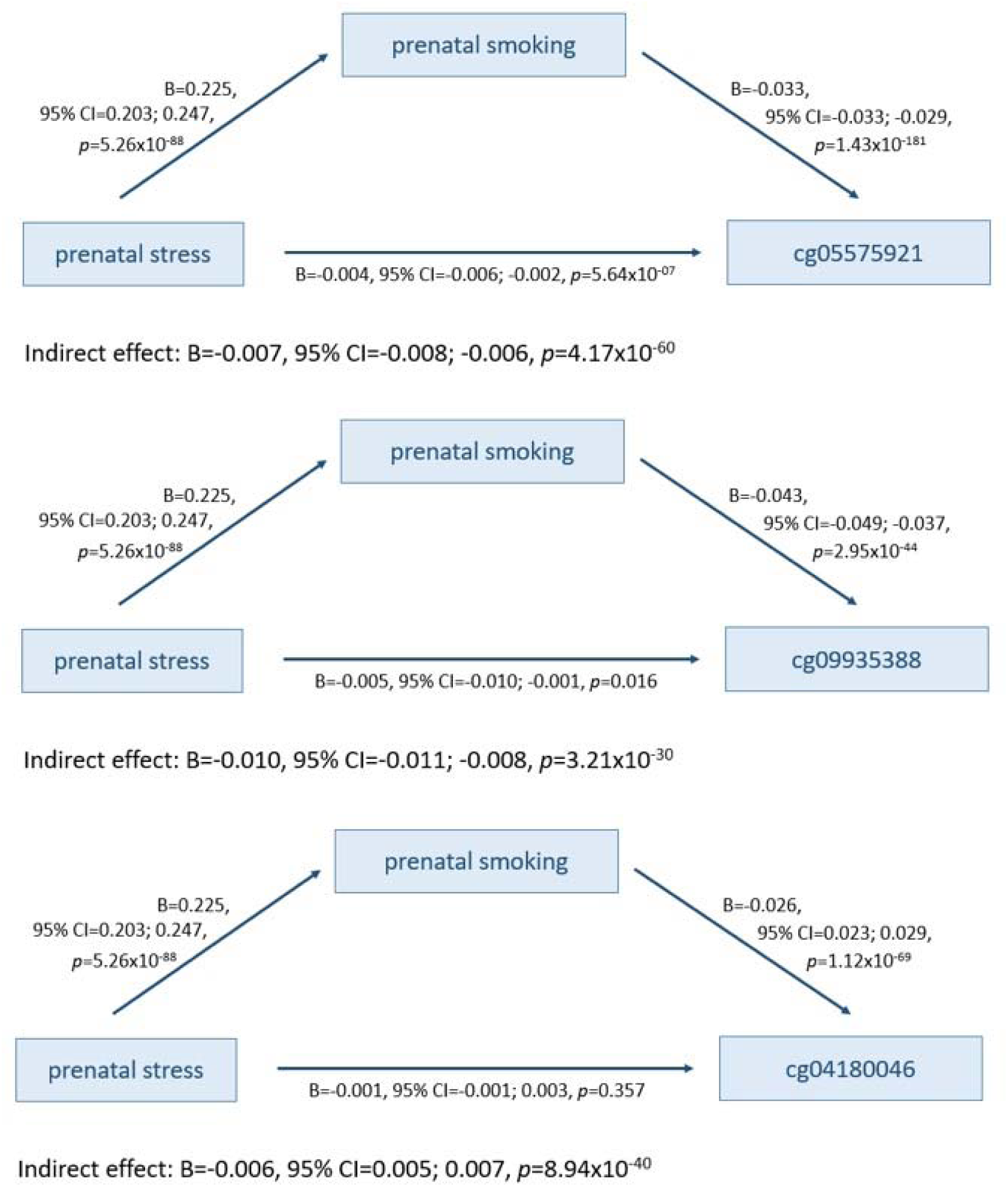
Mediation of cumulative prenatal stress associations with DNA methylation by maternal tobacco smoking during pregnancy (prenatal smoking)

### Pathway enrichments

A Gene Ontology analysis of 3,248 suggestive (*p*<5×10^-08^) SNPs in the GxEmodel yielded 145 overrepresented pathways and 12 underrepresented pathways (**Supplemental Table 4**). The overrepresented pathways were predominantly linked to neuronal development and synaptic transmission. The underrepresented pathways were linked, amongst others, to DNA repair processes. A Gene Ontology analysis of 2,613 suggestive CpGs in the GxEmodel yielded 35 overrepresented pathways (**Supplemental Table 5**), among which neuronal development-related pathways were predominant.

## Discussion

In this study, we investigated SNP-by-prenatal-stress interactions on DNAm at birth, for the first time at the genome- and epigenome-wide level. From the GxEmodel, we report five SNP-by-prenatal-stress interactions on DNAm after multiple testing correction, including five unique, of which two independent, SNPs in *CHD2* and *ORC5,* and two unique CpGs in *EPPK1*. By comparison, the Gmodel yielded 691,202 associations of SNPs and DNAm, including 181,133 unique SNPs (48% of investigated SNPs) and 54,809 unique CpGs (59% of investigated CpGs), and the Emodel identified three associations between cumulative prenatal stress and DNAm at CpGs in *AHRR, GFI1*, and *MYO1G*, which are known DNAm loci for smoking exposure. Together, these results do not support the notion that GxEmodels better predict DNAm than G- or E-models alone^12, 13, 55^.

Significant results for the GxEmodel were scarce, which might in part be explained by the stringent Bonferroni multiple testing correction in which analyses were considered as independent. Due to the scale of the analyses, this resulted in a very low *p*-value threshold. However, a Bonferroni threshold might be overly stringent as there was a correlational structure among SNPs and among CpGs, and we also tested the same SNP 86,236 times, and the same CpG 374,152 times. Using a suggestive (genome-wide) significance threshold, Gene Ontology analyses pointed to enrichment of neuronal development related pathways. Moreover, previous GWASs have associated variation in *ORC5*, in or near which four of the associated SNPs were located with entorhinal cortex volume^56^, an area important for memory processing, bordering the hippocampus and particularly rich in corticoid receptors^57^. These findings fit the developmental origins of health and disease perspective^58^, which poses that the prenatal environment programs organ structure and function (in this case in interaction with the offspring genotype), as well as findings from our own lab that cumulative prenatal stress is related to childhood subcortical brain volumes^16^, and that prenatal stress, *beyond* postnatal stress, predicts internalizing symptoms in childhood^15^.

In contrast to the GxEmodel, significant associations in the Gmodel were abundant, indicating that many common SNPs are involved in epigenetic programming and in turn, that DNAm is under strong genetic control. This confirms earlier meQTL studies, which also identified numerous genetic effects on the epigenome^21, 34^. The abundance of genetic effects and far fewer GxE associations are contrary to the notion put forward by other studies that gene-environment interaction studies perform better than studies of genetic main effects alone in terms of predicting DNA metylation^12, 13, 55^. The difference may lie in the different approach; in these studies of genetic interaction effects with multiple prenatal environments^12, 13^ and with adverse childhood experiences^55^, models were compared based on fit indices without adjustment for multiple testing, and results were found to be convergent between cohorts (e.g. GxEmodel to be superior over Gmodel or Emodel) for a CpG even if the SNP or environmental variable differed for that CpG. By contrast, we did not directly compare the different models, yet compare the amount of Bonferroni corrected significant results of each model type.

What emerges from our results, however, is that the GxEmodel yields *different* results than when looking at genetic or environmental main effects alone. Follow-up analyses showed that SNPs and CpGs brought forward by the GxEmodel were not more likely to have been identified as, or related to an meQTL. For future GxE studies on DNAm, this means that only testing GxE interactions among significant findings in the Gmodel^12^, would reduce the multiple testing burden, but might result in selective findings and may miss true GxE effects.

The Emodel yielded limited evidence of associations between cumulative prenatal stress and DNAm, which is in line with previous studies^9–11^. Moreover, whereas associations in the GxEmodel did not seem to be related to prenatal smoking and drinking behavior, gestational age or birthweight, the Emodel associations all could be largely explained by smoking behavior of the mother during pregnancy. Indeed, the look-up of related CpGs showed that these are top-hits in smoking EWASs^59–61^. Furthermore, whereas the GxEmodel results could not be explained by one of the types of stressors in particular, thereby ascribing to the notion that associations were due to the cumulative nature of prenatal stress rather than to the unique contribution of a specific stressor, contextual risk and interpersonal risk provided unique contributions to the results from the Emodel. These results fit with a recent EWAS meta-analysis of maternal educational attainment, often taken as an indicator of socio-economic position, which was also enriched for CpGs related to prenatal smoking^48^ as well as an EWAS on victimization stress in children, of which results could also largely be explained by smoking^62^. Taken together, we conclude that prenatal maternal stress associations with the offspring epigenome are not independent of maternal smoking behavior.

Results of this study should be interpreted in light of several limitations. First, effect sizes were small and Gene Ontology enrichment analysis of suggestive hits seemed to indicate that subthreshold findings are informative – hence larger sample sizes will likely be necessary to identify relevant gene-by-prenatal stress interactions with greater statistical power. However, to the best of our knowledge, this is the largest effort thus far to identify gene-by-prenatal stress effects on the epigenome. Moreover, we included a comprehensive measure of cumulative prenatal stress, capturing multiple domains of stress that often co-occur together – which has been uniquely harmonized between Generation R and ALSPAC, making it difficult to include a larger sample size at this time point. Second, there was heterogeneity between the (sub-)cohorts in the GxE association of a SNP in *CHD2* with a nearby CpG, which reduces the robustness of the finding. However, as the I^2^ measure that was used for heterogeneity is relatively sensitive, this does not necessarily mean the finding is a false positive. Future studies are needed to examine this association in more detail, preferably in multiple cohorts. Third, whereas SNPs and CpGs included in these models span the full genome, we reduced the number of probes based on intercorrelation and/or variability to minimize the burden of multiple testing. This does mean, however, that it is possible that we missed associations. Fourth, DNA methylation is tissue-specific and interactive effects of prenatal stress and offspring genotype on DNA methylation may differ between blood, which we used as an easily accessible tissue in population-based studies, and other, potentially more relevant tissues, such as brain – however, even in blood we found an epigenetic pattern of neurodevelopmental pathways. Fifth, the Generation R and ALSPAC are populations are generally selected towards being slightly healthier and more affluent than the general population, which may affect the generalizability of findings. It may also have reduced variation in prenatal stress and thereby the power to detect true associations. Also, as the epigenetic samples only include children of European ancestry, generalizability to populations of other ancestries may be limited. In the future, studies in populations of other ancestries are necessary to understand how genotype-by-prenatal-stress associates with DNAm at birth across different populations. Last, this is an observational study. Intervention studies on reducing prenatal stress^63–65^ might help understand the degree to which gene-by-prenatal stress associations are causal in nature. Furthermore, more research would be necessary to understand the consequences of genotype-by-prenatal-stress associations found. The enrichment analyses indicated that neuronal development might be involved, yet more research is necessary to understand for which aspects of neuronal development this would be the case and to what degree.

In conclusion, in this comprehensive study of genotype-by-prenatal stress interactions on DNAm, we report suggestive findings that cumulative prenatal stress interacts with the child’s genome on DNA methylation in or close to genes related to neuronal development. Importantly, these results do not support the idea that gene-environment interactions on the epigenome are more abundant than gene effects alone, as we found many more associations in our genetic main effect model than in our interactive model. In the future, larger studies and studies including participants of different genetic ancestries are needed to identify associations with smaller effect sizes and generate results that are more generalizable.

## Supporting information

Supplemental Results

## Data Availability

Data from the Generation R Study are available upon reasonable request to the director of the Generation R Study, subject to local, national and European rules and regulations. ALSPAC data access is through a system of managed open access. The ALSPAC access policy (http://www.bristol.ac.uk/media-library/sites/alspac/documents/researchers/data-access/ALSPAC_Access_Policy.pdf) describes the process of accessing the data and samples in detail, and outlines the costs associated with doing so.

## Acknowledgements

The Generation R Study is conducted by Erasmus MC, University Medical Center Rotterdam in close collaboration with the School of Law and Faculty of Social Sciences of the Erasmus University Rotterdam, the Municipal Health Service Rotterdam area, Rotterdam, the Rotterdam Homecare Foundation, Rotterdam and the Stichting Trombosedienst & Artsenlaboratorium Rijnmond (STAR-MDC), Rotterdam. We gratefully acknowledge the contribution of children and parents, general practitioners, hospitals, midwives and pharmacies in Rotterdam. The generation and management of the Illumina 450K methylation array data (EWAS data) for the Generation R Study was executed by the Human Genotyping Facility of the Genetic Laboratory of the Department of Internal Medicine, Erasmus MC, the Netherlands. We thank Mr. Michael Verbiest, Ms. Mila Jhamai, Ms. Sarah Hunter, Mr. Marijn Verkerk and Dr. Lisette Stolk for their help in creating the EWAS database. We thank Dr. A. Teumer for his work on the quality control and normalization scripts.

We are extremely grateful to all the families who took part in the ALSPAC study, the midwives for their help in recruiting them, and the whole ALSPAC team, which includes interviewers, computer and laboratory technicians, clerical workers, research scientists, volunteers, managers, receptionists and nurses.

## References

1. Hentges RF, Graham SA, Plamondon A, Tough S, Madigan S. A developmental cascade from prenatal stress to child internalizing and externalizing problems. Journal of pediatric psychology 2019; 44(9): 1057–1067.

2. Dancause KN, Laplante DP, Hart KJ, O’Hara MW, Elgbeili G, Brunet A, King S. Prenatal stress due to a natural disaster predicts adiposity in childhood: the Iowa Flood Study. Journal of obesity 2015; 2015.

3. Flanigan C, Sheikh A, DunnGalvin A, Brew BK, Almqvist C, Nwaru BI. Prenatal maternal psychosocial stress and offspring’s asthma and allergic disease: a systematic review and meta-analysis. Clinical & Experimental Allergy 2018; 48(4): 403–414.

4. Joubert BR, Felix JF, Yousefi P, Bakulski KM, Just AC, Breton C et al. DNA methylation in newborns and maternal smoking in pregnancy: genome-wide consortium meta-analysis. The American Journal of Human Genetics 2016; 98(4): 680–696.

5. Mulder RH, Walton E, Neumann A, Houtepen LC, Felix JF, Bakermans-Kranenburg MJ et al. Epigenomics of being bullied: changes in DNA methylation following bullying exposure. Epigenetics 2020: 1–15.

6. Vehmeijer FOL, Küpers LK, Sharp GC, Salas LA, Lent S, Jima DD et al. DNA methylation and body mass index from birth to adolescence: meta-analyses of epigenome-wide association studies. Genome medicine 2020; 12: 1–15.

7. Reese SE, Xu C-J, Herman T, Lee MK, Sikdar S, Ruiz-Arenas C et al. Epigenome-wide meta-analysis of DNA methylation and childhood asthma. Journal of Allergy and Clinical Immunology 2018.

8. Mulder RH, Rijlaarsdam J, Luijk MPCM, Verhulst FC, Felix JF, Tiemeier H et al. Methylation matters: FK506 binding protein 51 (FKBP5) methylation moderates the associations of FKBP5 genotype and resistant attachment with stress regulation. Development and Psychopathology 2017; 29(2): 491–503.

9. Sammallahti S, Hidalgo APC, Tuominen S, Malmberg A, Mulder RH, Brunst KJ et al. Maternal anxiety during pregnancy and newborn epigenome-wide DNA methylation. Molecular Psychiatry 2021: 1–14.

10. Kotsakis Ruehlmann A, Sammallahti S, Cortés Hidalgo AP, Bakulski KM, Binder EB, Campbell ML et al. Epigenome-wide meta-analysis of prenatal maternal stressful life events and newborn DNA methylation. Molecular Psychiatry 2023: 1–11.

11. Rijlaarsdam J, Pappa I, Walton E, Bakermans-Kranenburg MJ, Mileva-Seitz VR, Rippe RCA et al. An epigenome-wide association meta-analysis of prenatal maternal stress in neonates: A model approach for replication. Epigenetics 2016; 11(2): 140–149.

12. Teh AL, Pan H, Chen L, Ong ML, Dogra S, Wong J et al. The effect of genotype and in utero environment on interindividual variation in neonate DNA methylomes. Genome Res 2014; 24(7): 1064–1074.

13. Czamara D, Eraslan G, Page CM, Lahti J, Lahti-Pulkkinen M, Hämäläinen E et al. Integrated analysis of environmental and genetic influences on cord blood DNA methylation in new-borns. Nature Communications 2019; 10(1): 1–18.

14. Cecil CAM, Lysenko LJ, Jaffee SR, Pingault JB, Smith RG, Relton CL et al. Environmental risk, Oxytocin Receptor Gene (OXTR) methylation and youth callous-unemotional traits: a 13-year longitudinal study. Mol Psychiatry 2014.

15. Defina S, Woofenden T, Baltramonaityte V, Pariante CM, Lekadir K, Jaddoe VWV et al. Effects of Pre-and Postnatal Early-Life Stress on Internalizing, Adiposity and Their Comorbidity. Journal of the American Academy of Child & Adolescent Psychiatry 2023.

16. Bolhuis K, Mulder RH, de Mol CL, Defina S, Warrier V, White T et al. Mapping gene by early life stress interactions on child subcortical brain structures: A genome-wide prospective study. JCPP Advances 2022: e12113.

17. Kooijman MN, Kruithof CJ, van Duijn CM, Duijts L, Franco OH, van IJzendoorn MH et al. The Generation R Study: design and cohort update 2017. European journal of epidemiology 2016; 31(12): 1243–1264.

18. Fraser A, Macdonald-Wallis C, Tilling K, Boyd A, Golding J, Davey Smith G et al. Cohort profile: the Avon Longitudinal Study of Parents and Children: ALSPAC mothers cohort. International Journal of Epidemiology 2012; 42(1): 97–110.

19. Boyd A, Golding J, Macleod J, Lawlor DA, Fraser A, Henderson J et al. Cohort profile: the ‘children of the 90s’—the index offspring of the Avon Longitudinal Study of Parents and Children. International Journal of Epidemiology 2013; 42(1): 111–127.

20. Medina-Gomez C, Felix JF, Estrada K, Peters MJ, Herrera L, Kruithof CJ et al. Challenges in conducting genome-wide association studies in highly admixed multi-ethnic populations: the Generation R Study. European journal of epidemiology 2015; 30(4): 317–330.

21. Gaunt TR, Shihab HA, Hemani G, Min JL, Woodward G, Lyttleton O et al. Systematic identification of genetic influences on methylation across the human life course. Genome Biology 2016; 17(1): 61.

22. Purcell S, Neale B, Todd-Brown K, Thomas L, Ferreira MAR, Bender D et al. PLINK: a tool set for whole-genome association and population-based linkage analyses. The American journal of human genetics 2007; 81(3): 559–575.

23. Buuren Sv, Groothuis-Oudshoorn K. mice: Multivariate imputation by chained equations in R. Journal of statistical software 2010: 1–68.

24. R Core Team. R: A language and environment for statistical computing. 2013.

25. Lehne B, Drong AW, Loh M, Zhang W, Scott WR, Tan S-T et al. A coherent approach for analysis of the Illumina HumanMethylation450 BeadChip improves data quality and performance in epigenome-wide association studies. Genome Biology 2015; 16(1): 37.

26. Min JL, Hemani G, Davey Smith G, Relton C, Suderman M, Hancock J. Meffil: efficient normalization and analysis of very large DNA methylation datasets. Bioinformatics 2018.

27. Mulder RH, Neumann A, Cecil CAM, Walton E, Houtepen LC, Simpkin AJ et al. Epigenome-wide change and variation in DNA methylation in childhood: Trajectories from birth to late adolescence. Human Molecular Genetics 2021.

28. Chen Y-a, Choufani S, Grafodatskaya D, Butcher DT, Ferreira JC, Weksberg R. Cross-reactive DNA microarray probes lead to false discovery of autosomal sex-associated DNA methylation. The American Journal of Human Genetics 2012; 91(4): 762–764.

29. Gervin K, Salas LA, Bakulski KM, Van Zelm MC, Koestler DC, Wiencke JK et al. Systematic evaluation and validation of reference and library selection methods for deconvolution of cord blood DNA methylation data. Clinical epigenetics 2019; 11(1): 1–15.

30. Leek JT, Storey JD. Capturing heterogeneity in gene expression studies by surrogate variable analysis. PLoS genetics 2007; 3(9): e161.

31. Pan H, Holbrook JD, Karnani N, Kwoh CK. Gene, Environment and Methylation (GEM): a tool suite to efficiently navigate large scale epigenome wide association studies and integrate genotype and interaction between genotype and environment. BMC bioinformatics 2016; 17(1): 1–8.

32. Willer CJ, Li Y, Abecasis GR. METAL: fast and efficient meta-analysis of genomewide association scans. Bioinformatics 2010; 26(17): 2190–2191.

33. Chandler J, Cumpston M, Li T, Page MJ, Welch V. Cochrane handbook for systematic reviews of interventions. *Hoboken*: *Wiley* 2019.

34. Min JL, Hemani G, Hannon E, Dekkers KF, Castillo-Fernandez J, Luijk R et al. Genomic and phenotypic insights from an atlas of genetic effects on DNA methylation. Nature genetics 2021; 53(9): 1311–1321.

35. Battram T, Yousefi P, Crawford G, Prince C, Babaei MS, Sharp G et al. The EWAS Catalog: a database of epigenome-wide association studies. Wellcome open research 2022; 7.

36. Liu X, Tian D, Li C, Tang B, Wang Z, Zhang R et al. GWAS Atlas: an updated knowledgebase integrating more curated associations in plants and animals. Nucleic Acids Research 2023; 51(D1): D969–D976.

37. Wang K, Li M, Hakonarson H. ANNOVAR: functional annotation of genetic variants from high-throughput sequencing data. Nucleic acids research 2010; 38(16): e164–e164.

38. Rosseel Y. lavaan: An R package for structural equation modeling. Journal of statistical software 2012; 48: 1–36.

39. Grote S, Grote MS. Package ‘GOfuncR’. 2018.

40. Bohlin J, Håberg SE, Magnus P, Reese SE, Gjessing HK, Magnus MC et al. Prediction of gestational age based on genome-wide differentially methylated regions. Genome Biology 2016; 17(1): 207.

41. Li C, Gao W, Gao Y, Yu C, Lv J, Lv R et al. Age prediction of children and adolescents aged 6-17 years: an epigenome-wide analysis of DNA methylation. Aging (Albany NY) 2018; 10(5): 1015.

42. Islam SA, Goodman SJ, MacIsaac JL, Obradović J, Barr RG, Boyce WT, Kobor MS. Integration of DNA methylation patterns and genetic variation in human pediatric tissues help inform EWAS design and interpretation. Epigenetics & chromatin 2019; 12: 1–18.

43. Henneman P, Bouman A, Mul A, Knegt L, van der Kevie-Kersemaekers A-M, Zwaveling-Soonawala N et al. Widespread domain-like perturbations of DNA methylation in whole blood of Down syndrome neonates. PLoS One 2018; 13(3): e0194938.

44. Hillary RF, Ng HK, McCartney DL, Elliott HR, Walker RM, Campbell A et al. Blood-based epigenome-wide analyses of chronic low-grade inflammation across diverse population cohorts. Cell Genomics 2024; 4(5).

45. Dugué PA, Wilson R, Lehne B, Jayasekara H, Wang X, Jung CH et al. Alcohol consumption is associated with widespread changes in blood DNA methylation: analysis of cross-sectional and longitudinal data. Addiction Biology 2021; 26(1): e12855.

46. Liu C, Marioni RE, Hedman ÅK, Pfeiffer L, Tsai P-C, Reynolds LM et al. A DNA methylation biomarker of alcohol consumption. Molecular Psychiatry 2018; 23(2): 422–433.

47. Stephenson M, Bollepalli S, Cazaly E, Salvatore JE, Barr P, Rose RJ et al. Associations of alcohol consumption with epigenome-wide DNA methylation and epigenetic age acceleration: individual-level and co-twin comparison analyses. Alcoholism: Clinical and Experimental Research 2021; 45(2): 318–328.

48. Choudhary P, Monasso GS, Karhunen V, Ronkainen J, Mancano G, Howe CG et al. Maternal educational attainment in pregnancy and epigenome-wide DNA methylation changes in the offspring from birth until adolescence. Molecular Psychiatry 2024; 29(2): 348–358.

49. Bermingham ML, Walker RM, Marioni RE, Morris SW, Rawlik K, Zeng Y et al. Identification of novel differentially methylated sites with potential as clinical predictors of impaired respiratory function and COPD. EBioMedicine 2019; 43: 576–586.

50. Imboden M, Wielscher M, Rezwan FI, Amaral AFS, Schaffner E, Jeong A et al. Epigenome-wide association study of lung function level and its change. European Respiratory Journal 2019; 54(1).

51. Carmona JJ, Barfield RT, Panni T, Nwanaji-Enwerem JC, Just AC, Hutchinson JN et al. Metastable DNA methylation sites associated with longitudinal lung function decline and aging in humans: an epigenome-wide study in the NAS and KORA cohorts. Epigenetics 2018; 13(10-11): 1039–1055.

52. Terzikhan N, Xu H, Edris A, Bracke KR, Verhamme FM, Stricker BHC et al. Epigenome-wide association study on diffusing capacity of the lung. ERJ Open Research 2021; 7(1).

53. Logue MW, Miller MW, Wolf EJ, Huber BR, Morrison FG, Zhou Z et al. An epigenome-wide association study of posttraumatic stress disorder in US veterans implicates several new DNA methylation loci. Clinical epigenetics 2020; 12: 1–14.

54. Smith AK, Ratanatharathorn A, Maihofer AX, Naviaux RK, Aiello AE, Amstadter AB et al. Epigenome-wide meta-analysis of PTSD across 10 military and civilian cohorts identifies methylation changes in AHRR. Nature Communications 2020; 11(1): 5965.

55. Czamara D, Tissink E, Tuhkanen J, Martins J, Awaloff Y, Drake AJ et al. Combined effects of genotype and childhood adversity shape variability of DNA methylation across age. Translational Psychiatry 2021; 11(1): 88.

56. Zhao B, Luo T, Li T, Li Y, Zhang J, Shan Y et al. Genome-wide association analysis of 19,629 individuals identifies variants influencing regional brain volumes and refines their genetic co-architecture with cognitive and mental health traits. Nature genetics 2019; 51(11): 1637–1644.

57. Lupien SJ, Maheu F, Tu M, Fiocco A, Schramek TE. The effects of stress and stress hormones on human cognition: Implications for the field of brain and cognition. Brain and cognition 2007; 65(3): 209–237.

58. Barker DJ. The fetal and infant origins of adult disease. BMJ: British Medical Journal 1990; 301(6761): 1111.

59. Andersen AM, Philibert RA, Gibbons FX, Simons RL, Long J. Accuracy and utility of an epigenetic biomarker for smoking in populations with varying rates of false self-report. American Journal of Medical Genetics Part B: Neuropsychiatric Genetics 2017; 174(6): 641–650.

60. Philibert R, Hollenbeck N, Andersen E, McElroy S, Wilson S, Vercande K et al. Reversion of AHRR demethylation is a quantitative biomarker of smoking cessation. Frontiers in psychiatry 2016; 7: 55.

61. Bojesen SE, Timpson N, Relton C, Smith GD, Nordestgaard BG. AHRR (cg05575921) hypomethylation marks smoking behaviour, morbidity and mortality. Thorax 2017; 72(7): 646–653.

62. Marzi SJ, Sugden K, Arseneault L, Belsky DW, Burrage J, Corcoran DL et al. Analysis of DNA methylation in young people: limited evidence for an association between victimization stress and epigenetic variation in blood. American Journal of Psychiatry 2018; 175(6): 517–529.

63. Virgili M. Mindfulness-based interventions reduce psychological distress in working adults: a meta-analysis of intervention studies. Mindfulness 2015; 6(2): 326–337.

64. Turner K, McCarthy VL. Stress and anxiety among nursing students: A review of intervention strategies in literature between 2009 and 2015. Nurse Education in Practice 2017; 22: 21–29.

65. Yusufov M, Nicoloro-SantaBarbara J, Grey NE, Moyer A, Lobel M. Meta-analytic evaluation of stress reduction interventions for undergraduate and graduate students. International Journal of Stress Management 2019; 26(2): 132.

