## Supplemental Information for "Interactive effects of genotype with prenatal stress on DNA methylation at birth"

In Generation R, 9,778 pregnant mothers gave birth to 9,749 live-born children. Genotype data were available for 7,502 children and for 5,684 of these, data on cumulative prenatal stress were also available. A total of 1,264 of these children had DNAm data at birth measured with the Illumina Infinium HumanMethylation450 BeadChip (Illumina Inc., San Diego, CA) (*GENR 450K*), all of which had genetic European ancestry (<4 SD from Hapmap II European descent [CEU] panel mean value for the first four principal components). Of these, 33 children were genotyped using the GSA-MD v2.0 array (Illumina Inc., CA, USA) and could not be imputed jointly with the rest of the sample due to little SNP-overlap. Since this group was too small to analyze separately, these children were excluded. Last, children with cryptic relatedness (IBD>0.15) were removed, one of each related pair based on data availability or otherwise randomly, leaving a final sample of 1,224 children in *GENR 450K*.

Secondly, from the group of 5,684 children with genotype and cumulative prenatal stress data in Generation R, another set of 968 (non-overlapping) children had DNAm data at birth measured with the Illumina Infinium MethylationEPIC v1.0 Beadchip (*GENR EPIC*). Children with cryptic relatedness (identity by descent [IBD]>0.15) were removed, one of each related pair based on data availability or otherwise randomly, leaving 949 children in *GENR EPIC*.

In the ALSPAC study, the initial number of pregnancies enrolled was 14,541. Of these, 13,988 children were alive at 1 year of age. When the oldest children were approximately 7 years of age, an attempt was made to bolster the initial sample with eligible cases who had failed to join the study originally. The phases of enrolment are described in more detail elsewhere (Boyd et al., 2013; Fraser et al., 2013). The total sample size for analyses using any data collected after the age of seven is therefore 15,447 pregnancies, resulting in 15,658 fetuses. Of these, 14,901 children were alive at 1 year of age. Of the original 14,541 initial pregnancies, 338 were from a woman who had already enrolled with a previous pregnancy, meaning 14,203 unique mothers were initially enrolled in the study. As a result of the additional phases of recruitment, a further 630 women who did not enroll originally have provided data since their child was 7 years of age. This provides a total of 14,833 unique women (G0mothers) enrolled in ALSPAC as of September 2021. Genotype data were available for 8,797 children and for 7,483 of these, data on their mother's cumulative prenatal stress were also available. DNAm data at birth measured with the Illumina Infinium HumanMethylation450 BeadChip (*ALSPAC 450K*) were available for 793 children as part of the Accessible Resource for Integrated Epigenomic Studies (ARIES) (Relton et al., 2015). Of these, 3 children with cryptic relatedness (IBD>0.15) were removed, one of each related pair based on data availability or otherwise randomly, leaving a final sample of 790 children in *ALSPAC 450K*.

**References**

Boyd, A., Golding, J., Macleod, J., Lawlor, D. A., Fraser, A., Henderson, J., . . . Davey Smith, G. (2013). Cohort profile: The 'Children of the 90s'-The index offspring of the avon longitudinal study of parents and children. *Int J Epidemiol, 42*(1), 111-127. doi:10.1093/ije/dys064

Fraser, A., Macdonald-Wallis, C., Tilling, K., Boyd, A., Golding, J., Davey Smith, G., . . . Lawlor, D. A. (2013). Cohort Profile: the Avon Longitudinal Study of Parents and Children: ALSPAC mothers cohort. *Int. J. Epidemiol., 42*(1), 97-110.

Relton, C. L., Gaunt, T., McArdle, W., Ho, K., Duggirala, A., Shihab, H., . . . Reik, W. (2015). Data resource profile: accessible resource for integrated epigenomic studies (ARIES). *Int. J. Epidemiol., 44*(4), 1181-1190.
