## Supplemental Figures for "Interactive effects of genotype with prenatal stress on DNA methylation at birth"

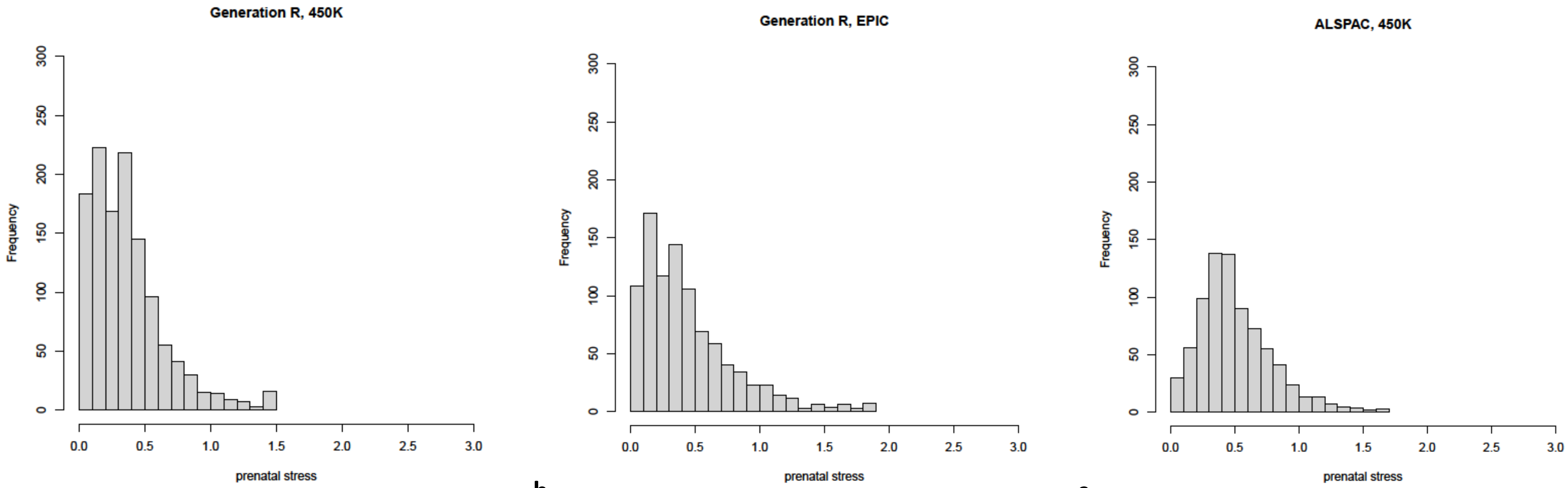


**a.** **b.** **c.**

**Supplemental Figure 1.** Histograms of prenatal maternal stress in the **a)** *GENR 450K* subcohort, **b)** *GENR EPIC* subcohort, and **c)** *ALSPAC 450K* cohort.

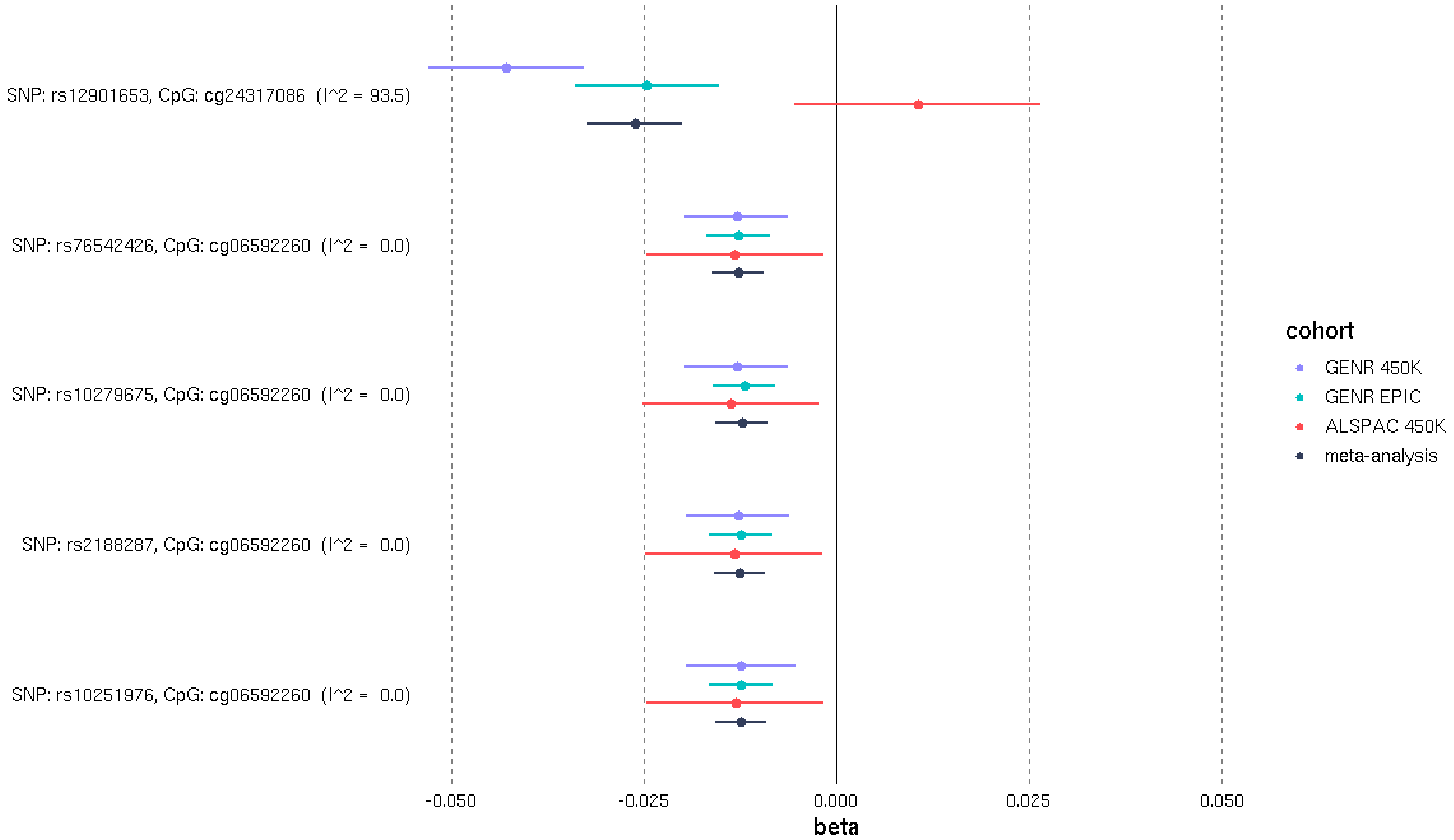


**Supplemental Figure 2.** Forest plot for significant (*p*<$1.55 \times 10^{-12}$) GxEmodel results of SNP by cumulative prenatal stress interaction associations with DNA methylation

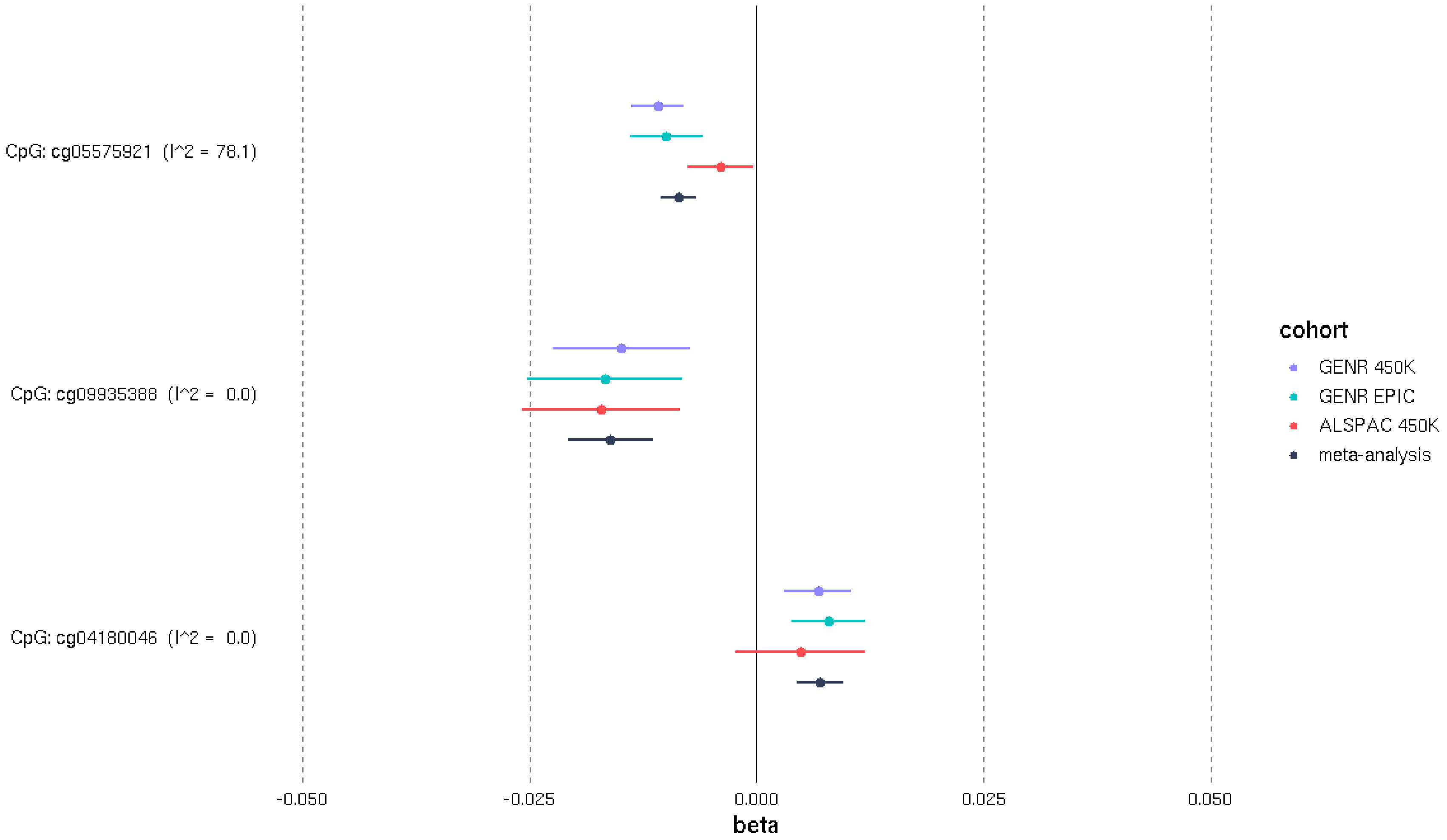


**Supplemental Figure 3.** Forest plot for significant (*p*<5.80x10$^{-07}$) Emodel results of cumulative prenatal stress associations with DNA methylation

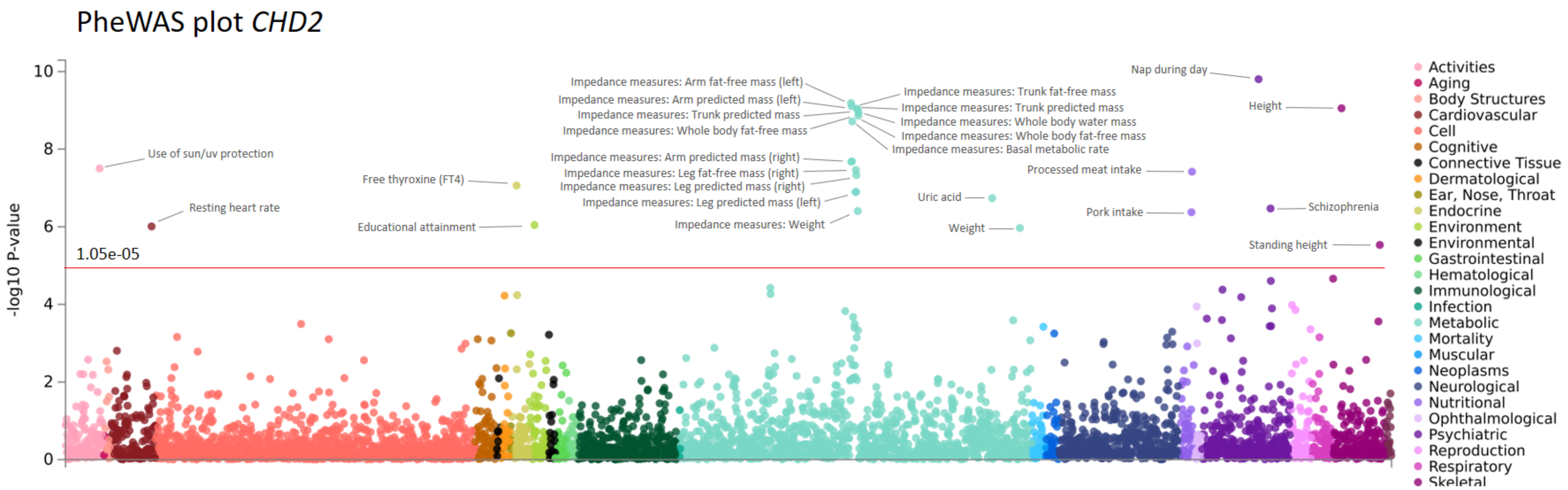


**Supplemental Figure 4.** Phenome-wide association study for *CHD2*, including 4756 GWASs, performed with the GWAS Atlas (https://atlas.ctglab.nl/PheWAS)

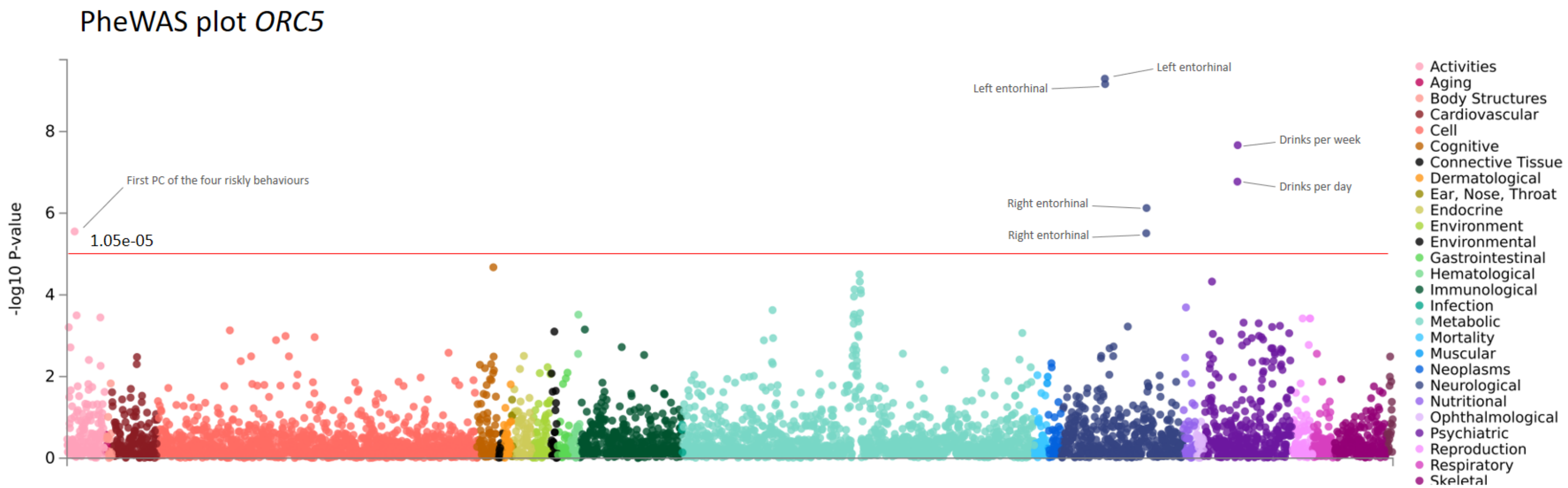


**Supplemental Figure 5.** Phenome-wide association study for *ORC5*, including 4756 GWASs, performed with the GWAS Atlas (https://atlas.ctglab.nl/PheWAS)

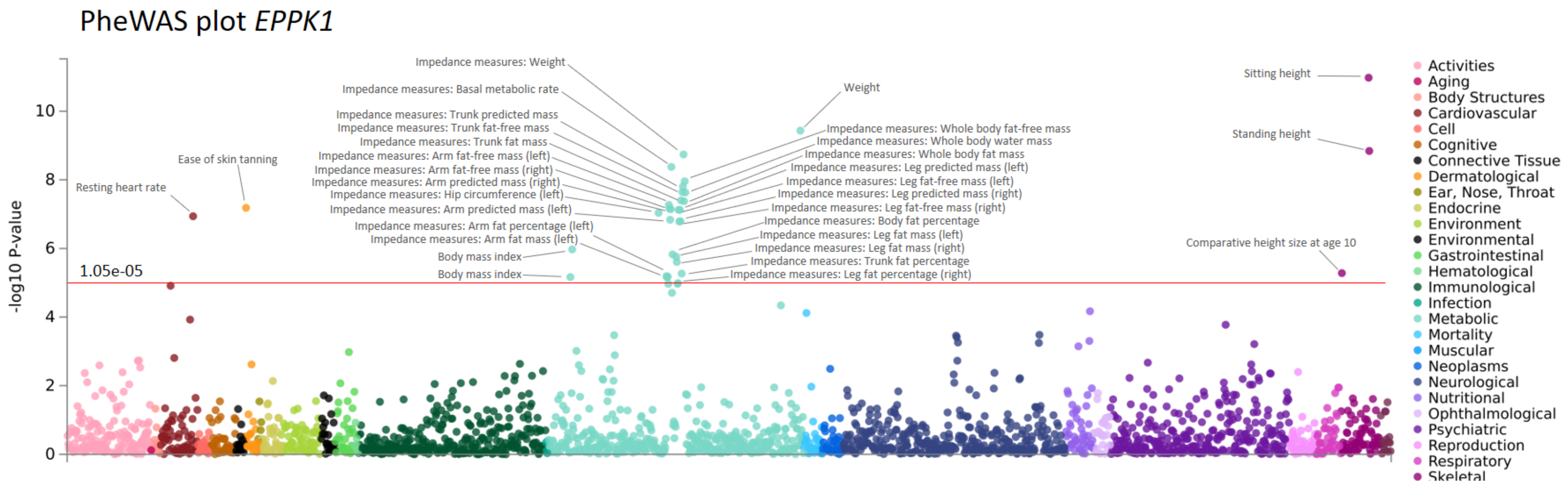


**Supplemental Figure 6.** Phenome-wide association study for *EPPK1*, including 4756 GWASs, performed with the GWAS Atlas (https://atlas.ctglab.nl/PheWAS)

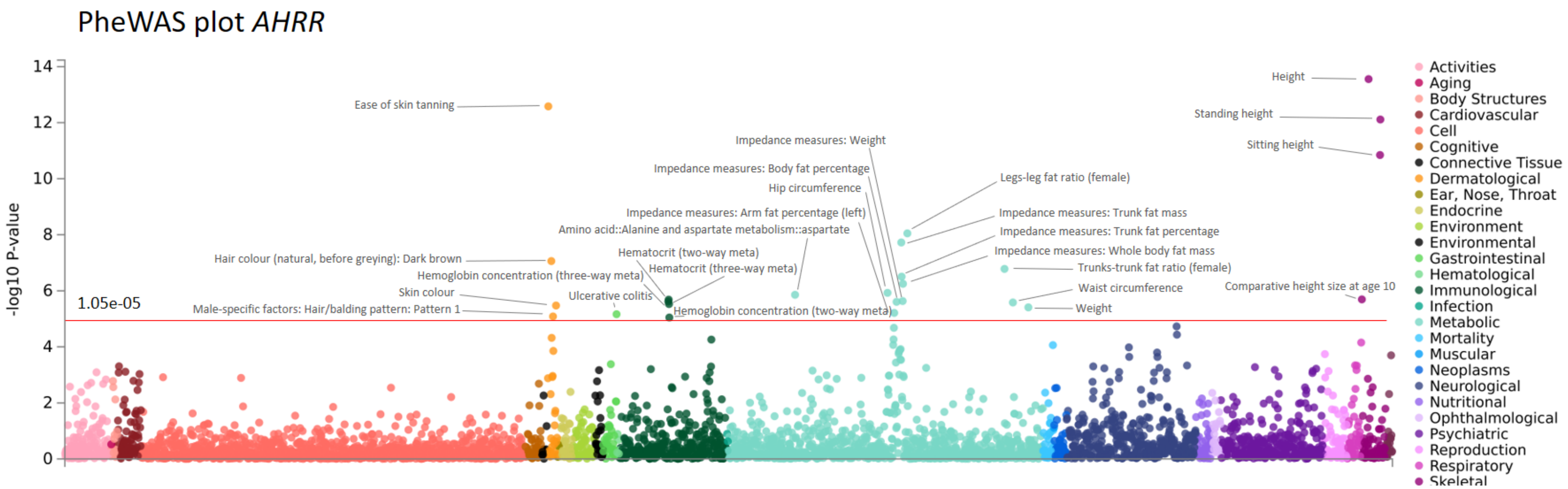


**Supplemental Figure 7.** Phenome-wide association study for *AHRR*, including 4756 GWASs, performed with the GWAS Atlas (https://atlas.ctglab.nl/PheWAS)

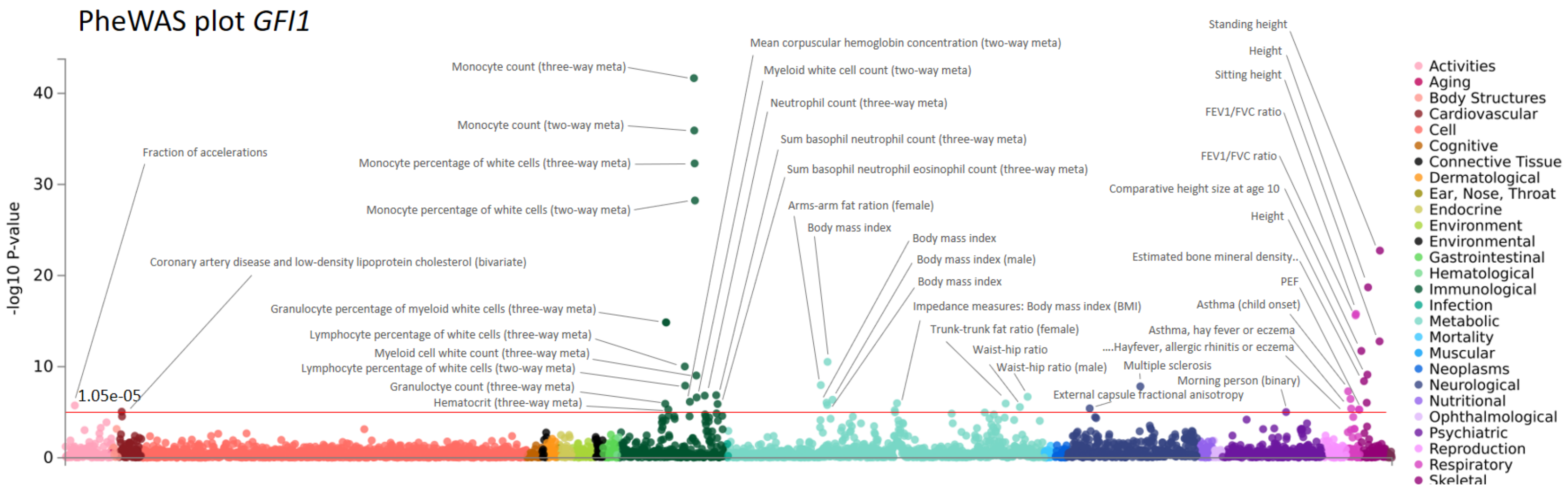


**Supplemental Figure 8.** Phenome-wide association study for *GFI1*, including 4756 GWASs, performed with the GWAS Atlas (https://atlas.ctglab.nl/PheWAS)

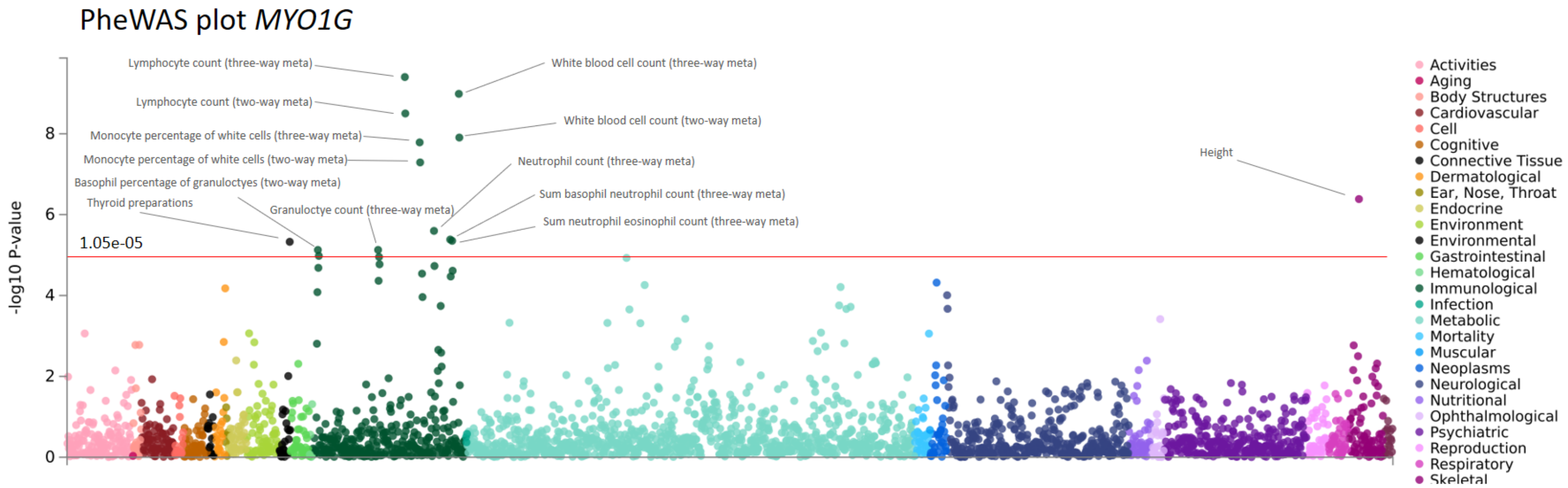


**Supplemental Figure 9.** Phenome-wide association study for *MYO1G*, including 4756 GWASs, performed with the GWAS Atlas (https://atlas.ctglab.nl/PheWAS)
