## Supplemental Results for "Interactive effects of genotype with prenatal stress on DNA methylation at birth"

**Supplemental Results S1**

Enrichment of known methylation quantitative trait loci in GxEmodel and Gmodel

Suggestive SNPs and CpGs in the GxEmodel ($p<5x10^{-08}$; 3,248 SNPs and 2,613 CpGs) and Gmodel (223,254 SNPs and 62,826 CpGs) were followed up to test enrichment for previously identified as (SNPs) or linked to (CpGs) meQTL*s* [1]. Among suggestive findings in the GxEmodel, SNPs were as likely to have been identified as an meQTL previously (4.6%) as other (non-suggestive) SNPs were (5.2%; OR=0.9 [95% CI=0.7-1.0], $p$=0.14), and suggestive CpGs were *less* likely to have been linked to an meQTL previously than other CpGs were (45% vs 50%; OR=0.8 [95% CI=0.8-0.9], $p=4x10^{-06}$).

By comparison, suggestive SNPs (i.e. suggestive meQTLs) in the Gmodel were more likely to already have been identified as an meQTL (8%) than other SNPs were (2%; OR=4.6 [95% CI=4.4-4.7], $p<2.23x10^{-308}$), as were suggestive CpGs (54% vs 40%; OR=1.8 [95% CI=1.7-1.8], $p=6.27x10^{-303}$).

**References**


1. Min JL, Hemani G, Hannon E, Dekkers KF, Castillo-Fernandez J, Luijk R *et al.* Genomic and phenotypic insights from an atlas of genetic effects on DNA methylation. *Nature genetics* 2021; **53**(9)**:** 1311-1321.
